# Improving on polygenic scores across complex traits using select and shrink with summary statistics

**DOI:** 10.1101/2022.09.13.22278911

**Authors:** J.P Tyrer, P. Peng, A.A DeVries, S.A Gayther, M.R Jones, P.D Pharoah

## Abstract

**Motivation:** As precision medicine advances, polygenic scores (PGS) have become increasingly important for clinical risk assessment. Many methods have been developed to create polygenic models with increased accuracy for risk prediction. Our select and shrink with summary statistics (S4) PGS method extends a previous method (polygenic risk score – continuous shrinkage (PRS-CS)) by using a continuous shrinkage prior on effect sizes with a selection strategy for including SNPs to create the best performing model.

**Results:** The S4 method provides overall improved PGS accuracy for UK Biobank participants when compared to LDpred2 and PRS-CS across a variety of phenotypes with differing genetic architectures. Additionally, the S4 method has higher estimated PGS accuracy over LDpred2 in Finnish and Japanese populations. Thus, the S4 method represents an improvement in overall PGS accuracy across multiple phenotypes and increases the transferability of PGS across ancestries.

**Availability and Implementation:** The S4 program is freely available at https://github.com/jpt34/S4_programs.

**Supplementary information:** Supplementary data [will be] available at *Bioinformatics* online.

## Introduction

Genome wide association studies (GWAS) have identified associations between common genetic variants and more than 3,300 phenotypes (Watanabe *et al*., 2019), revealing the highly polygenic architecture of many common traits. Polygenic scores (PGS) are a weighted sum of the known genome-wide risk alleles for a specific phenotype as calculated for an individual. Generally, the summary statistics of a genome-wide association study (GWAS) inform the selection and weighting of the common variants in a polygenic model used to calculate a PGS for an individual. Each GWAS variant confers only small risks individually, but their combined effects, when summarized as a PGS, may be substantial. As personalized medicine becomes a larger part of medical care, PGS may be clinically useful to help early detection, individual stratification, and prevention in the general population for a variety of diseases (Khera *et al*., 2018; Mavaddat *et al*., 2019; Chatterjee *et al*., 2016).

The development of novel methods for PGS estimation allow for different approaches to inform selection of variants and weighting of alleles. In creating a PGS method, it is important that a model is both accurate and computationally feasible. Variants are generally selected by the confidence of association and weighted by their effect on risk as determined from GWAS summary statistics (Wray *et al*., 2007; International Schizophrenia Consortium *et al*., 2009). Additionally, risk variants in high linkage disequilibrium (LD) with each other are often pruned or down-weighted to limit overrepresentation of highly correlated variants from the same association signal (International Schizophrenia Consortium *et al*., 2009). For both the GWAS summary statistics and LD reference panel, an ancestry matched cohort is ideal to improve accuracy. Additionally, for clinical usage, selection of fewer variants in the model will improve computational efficiency.

Previously, we presented our “select and shrink with summary statistics” (S4) PGS method which accurately and efficiently predicted the polygenic risk of epithelial ovarian cancer (Dareng *et al*., 2022). In this paper, we further demonstrate the accuracy of the S4 PGS method in risk predictions of multiple phenotypes, and compare the S4 PGS method to LDpred2 and PRS-CS, two other commonly used PGS methods (Dareng *et al*., 2022; Vilhjálmsson *et al*., 2015; Privé *et al*., 2020; Ge *et al*., 2019). LDpred is a Bayesian method that uses both a point-normal mixture distribution prior and LD information from a reference panel to estimate posterior mean causal effect sizes to improve accuracy in the PGS (Vilhjálmsson *et al*., 2015). LDpred2 improves the computational efficiency of LDpred, as well as its accuracy when causal variants are in long-range LD regions or are only a small proportion of the total variation (Privé *et al*., 2020). PRS-CS is another Bayesian method that uses a continuous shrinkage (CS) prior on effect sizes to accommodate a wide variety of genetic architectures while improving computational feasibility compared to LDpred (Ge *et al*., 2019). The S4 method uses a continuous shrinkage prior on effect sizes similar to PRS-CS, but also allows for improved penalization of rare SNPs by correcting for standard deviation of the estimate (Dareng *et al*., 2022).

Here, we first compare all three methods across diverse phenotypes for which summary GWAS results data were available and genotype level data for the same phenotype were available in UK Biobank (Sudlow *et al*., 2015). Across multiple phenotypes with varying genetic architectures, we find that the S4 method provides overall improved PGS accuracy. We also assess estimated accuracy of PGS derived using S4 and LDpred2 in Finnish and Japanese populations, and demonstrate that the S4 method creates a PGS with greater transferability across ancestries (Nagai *et al*., 2017; Kurki *et al*., 2022). We find that the computationally efficient S4 PGS method has strong potential for development of accurate PGS across a variety of phenotypes and populations.

## Methods

### Phenotypes and study populations

We performed PGS modeling and association testing for 12 phenotypes: Asthma, body mass index (BMI), breast cancer, coronary artery disease, endometrial cancer, height, inflammatory bowel disease (IBD), major depressive disorder, prostate cancer, schizophrenia, type 1 diabetes, and type 2 diabetes. These phenotypes were chosen to represent a variety of traits, and to include those influenced by epidemiological as well as genetic risk factors.

Published GWAS summary statistics were collected and used as input to form a polygenic model for each trait (Figure 1, Supplementary Table 1) (Demenais *et al*., 2018; Locke *et al*., 2015; Michailidou *et al*., 2017; Nikpay *et al*., 2015; Chen *et al*., 2016; Wood *et al*., 2014; de Lange *et al*., 2017; Wray *et al*., 2018; Schumacher *et al*., 2018; Schizophrenia Psychiatric Genome-Wide Association Study (GWAS) Consortium, 2011; Censin *et al*., 2017; Scott *et al*., 2017). We collected genotype and phenotype data and GWAS summary statistics from the UK Biobank (Sudlow *et al*., 2015), and only GWAS summary statistics from FinnGen (Kurki *et al*., 2022) and BioBank Japan (Nagai *et al*., 2017). The numbers of case and control samples used in each phenotype are detailed in Supplementary Table 1.

**Figure 1:**
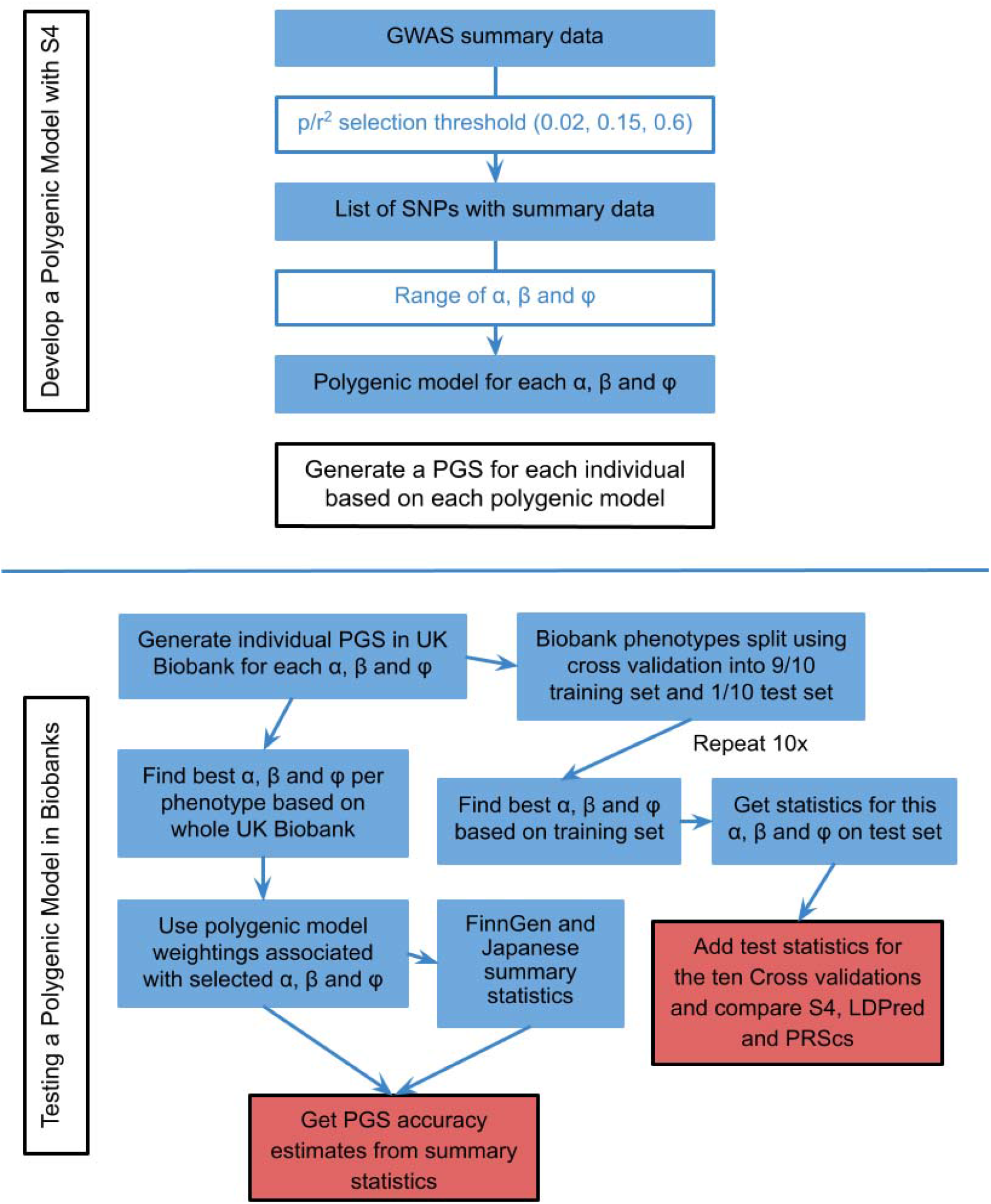
S4 Development and Testing Methods. Steps to develop each S4 polygenic model and testing accuracy of the S4 PGS’s in biobanks. The S4 method was compared against LDpred2 and PRS-CS.

### PGS models training and validation

The S4 PGS method was compared with LDPred2 and PRS-CS (Privé *et al*., 2020; Ge *et al*., 2019). To maintain a fair comparison, polygenic scores for all three methods were created as a linear function of 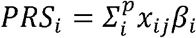, where *x*_*ij*_ represents the *i*th individual for the *j*th SNP out of *p* SNPs on an additive log scale, and *β*_*i*_ represents the weight, i.e. the log of the odds ratio, of the *j*th SNP. Genotypes are denoted as *x*, taking on the minor allele dosages of 0, 1 and 2. The three methods used different approaches to select and derive the optimal weights *β*_*i*_, which we described below.

We first prepared LD matrices (a set of between SNP pairwise correlations or r^2^) from the Oncoarray genotyping panel for breast, prostate and endometrial cancer and from an Illumina 610 genotyping panel for the other phenotypes (Amos *et al*., 2017; Pharoah *et al*., 2013; Phelan *et al*., 2017). The Oncoarray panel was chosen as the reference for the cancer phenotypes as the bulk of the samples used in the cancer summary statistics were genotyped on the OncoArray while the Illumina 610 panel is likely to match the non-cancer phenotypes more closely.

The S4 PGS method has been previously described in Dareng et al. 2022 (Dareng *et al*., 2022). We briefly review the main ideas of S4 PGS here. The S4 method first selects SNPs from the GWAS summary statistics to include in the model based on defined thresholds for the p-value/r^2^ ratio Figure 1). Each top GWAS SNP is iteratively added if the correlation with all the other SNPs included is less than 0.85. When the p-value divided by the correlation to other SNPs in the model is less than a specified threshold (0.02, 0.15, and 0.6 were tested in this analysis) no more SNPs were chosen (Table 3). At each threshold, the number of SNPs varied depending on phenotype and density of summary statistics coverage. The threshold of p/r^2^ < 0.6 was not tested on certain phenotypes when computationally infeasible. For BMI, rather than p/r^2^ < 0.6, all SNPs were included as the number of SNPs was still computationally feasible.

The weight for each of the selected SNPs was calculated using a continuous shrinkage prior which is applied according to tuning parameters □, β and □. □ controls the shrinkage of effect estimates around 0, β controls the shrinkage of larger effect estimates, and □ is the overall shrinkage parameter. □ was set to 0.1 and β was set to a range of values from 0.7 to 100 with increments of 0.1 up to 2.0, increments of 0.2 to 4.0, and eight more increments up to the maximum of 100. The values for □ were taken as 1, 1.2, 1.5, 2, 2.5, 3, 4, 5, 6, 8 multiplied by factors of 10 from 1e-10 to 1e10. The range of values was selected such that the probability of an absolute effect of at least 0.01 was reasonable, so that a different range of values was selected depending on the value of beta.

LDpred2 is a popular method for creating a PGS that estimates effect sizes with point-normal prior and a Gibbs sampler. The parameters selected to test the LDpred2 model followed the suggestions in the LDpred2 paper (Privé *et al*., 2020). For some of the phenotypes, the best h^2^ coefficient was outside the range suggested within the paper, so extra h^2^ values were generated to select the best fitting model. The aim was to make sure the best fitting model was not an extreme value of h^2^ (either biggest or smallest). LDpred2 assumes that a spike and slab prior fits the data well.

The PRS-CS model is also a Bayesian method that applies a continuous shrinkage prior on effect sizes. This model does not use the same SNP selection method as S4, which is able to better penalize rarer SNPs. For PRS-CS, the tuning parameter representing global shrinkage, □, was tested at 1e-6, 3e-6, 1e-5, 3e-5 etc. so that the best fitting value lied in the interior of this set of values. This ensures that the best fitting value is close to the global optimum.

We then used 10 fold cross-validation on the UK Biobank data to select the best tuning parameters for each phenotype (Sudlow *et al*., 2015). For each fold of the data, the PGS based on the SNP-specific weights corresponding to each combination of the tuning parameters was calculated and the performance of the PGS was assessed. The PGS that performed best was then tested in the remaining 10% of the data. Finally the best tuning parameters were selected by the PGS with the highest average prediction accuracy over the 10 folds, as calculated by area under the receiver operator curve (AUC) for categorical variables and correlation for continuous variables. For a comprehensive performance evaluation, besides AUC, we also reported odds ratio (OR) normalized by one standard deviation of PGS, and 95% confidence interval (CI).

### PGS performance evaluation in alternate populations

To test transferability to non-European populations, the best PGS models trained in UK Biobank for both the S4 method and LDpred2 method were evaluated in Finnish (FinnGen (Kurki *et al*., 2022)) and Japanese (BioBank Japan (Nagai *et al*., 2017)) cohorts. As no individual level genotypes are available for these cohorts, we estimated the PGS effects from the summary statistics and variants correlation matrix instead, as follows:

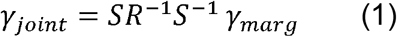

where *γ*_*joint*_ is the joint distribution (i.e. PGS effect), *γ*_*marg*_ is the marginal effects (i.e. summary statistics in the test set), *R* denotes the correlation matrix between the variants in the PGS model, and *S* represents the diagonal matrix of standard errors of the marginal effect estimates (i.e., the standard errors in the summary statistics). The standard errors of the marginal effects are inversely proportional to the standard deviation of the variants. Note that this derivation of the PGS effect estimate is similar to the PGS effect estimate delineated in LDPred2 (Privé *et al*., 2020), while the diagonal matrix *S* used in LDPred2 is based on the standard deviations of variants.

The regular calculation of PGS takes a weighted sum of minor allele dosages:*PGS*_*j*_ = Σ_*i*_*w*_*i*_ *x*_*ij*_ where *j* is the individual, *i* is the *i*th variant, *w*_*i*_ is the weight given to the *i*th SNP and *x*_*ij*_ is the dosage value of the *j*th individual at the *i*th variant. Therefore, the expected value of PGS can be written as:

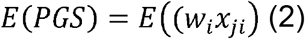

Consider this as a meta-analysis:

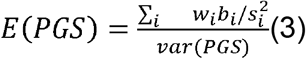

where *b*_*i*_ is the estimate effect for variant *i, s*_*i*_ is the standard error of variant *i, w*_*i*_ is the weight for the PGS, and *var(PGS)* is the variance for the PGS. However, the variance here is different from a normal meta-analysis as the variants are correlated (Lin and Sullivan, 2009). The variance is:

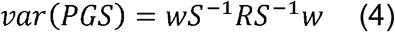

If R was the identity matrix, then this would reduce to a normal meta-analysis, even though analyzed on the same data, as then the variants would all be uncorrelated.

Therefore, we obtain the PGS coefficient, *PRS*_*b*_, and its standard error, *PRS*_*se*_, as follows:

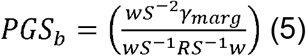

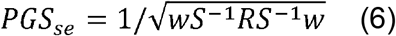

For calculating the estimated coefficient per standard deviation, we need to estimate the standard deviation of the PGS in the tested dataset. This is done by calculating 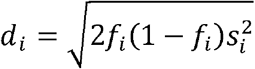 for each variant i where f_i_ is the frequency and s_i_ is the standard error of the beta estimate. As the variance of each variant will be smaller than 2*f*_*i*_(1−*f*_*i*_) because of imputation accuracy one of the lower values of d (0.2% percentile) is selected as the estimate for the population. Then:

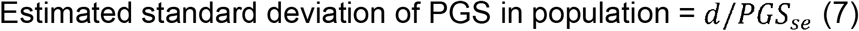

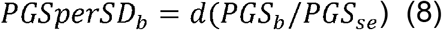

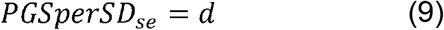

## Results

### S4 polygenic predictions of multiple phenotypes in UK Biobank

We applied the S4 PGS method to predict ten complex diseases (asthma, breast cancer, coronary artery disease, endometrial cancer, inflammatory bowel disease, major depressive disorder, prostate cancer, schizophrenia, type 1 diabetes, and type 2 diabetes), and two quantitative traits (body mass index and height) in the UK Biobank (Sudlow *et al*., 2015). GWAS summary statistics and individual level genotype data for each disease and trait were used to evaluate the performance of each S4 model. The optimal parameters and performance of best-fitting models for each phenotype are shown in Table 1. Prediction performance metrics included area under the receiver operating characteristic curve (AUC), odds ratio (OR) normalized by one standard deviation of PGS, and 95% confidence interval (CI). Among the 12 phenotypes, the AUC values of S4 PGS predictions ranged from 0.56 to 0.79, with better predictions in type 1 diabetes (AUC=0.79), inflammatory bowel disease (AUC=0.73), and schizophrenia (AUC=0.72). PGS associations were accessed by normalized OR, ranging from 0.22 for major depressive disorder to 1.14 for type 1 diabetes. The number of SNPs selected varied among phenotypes, ranging from 19,584 for type 2 diabetes to 1,239,271 for schizophrenia.

### S4 PGS outperforms other polygenic score prediction methods

We compared the S4 PGS method with LDPred2 (Privé *et al*., 2020) and PRS-CS (Ge *et al*., 2019), which had been shown to reliably predict polygenic scores on multiple phenotypes. Overall, the best S4 model performs better than LDPred2 in nine out of 11 phenotypes (Table 2). The S4 PGS method had better prediction accuracy and association for endometrial cancer (AUC=0.697, OR=0.396) and prostate cancer (AUC=0.710, OR=0.816) than LDPred2 (endometrial cancer AUC=0.597, OR=0.351; prostate cancer AUC=0.695, OR=0.755). The S4 models for asthma and height did not out-perform LDPred2. The comparisons between models were based on 10-fold cross-validation performance for both methods. We note that S4 PGS AUC values from cross-validations were similar to the AUC values derived from full training datasets (Table 1), indicating overfitting problems were less likely to occur during our model training. Consistent with results from previous S4 PGS predictions on epithelial ovarian cancer (Dareng *et al*., 2022), the S4 PGS methods used noticeably less SNPs than LDPred2 on all phenotypes except schizophrenia. The greatest difference was in type 1 diabetes, where LDPred2 (n=515,920) had 26-fold increased number of SNPs than S4 PGS (n=19,584).

We also analyzed the 10-fold cross-validation performance of PRS-CS on the same set of phenotypes. S4 PGS outperforms PRS-CS in all phenotypes tested (Supplementary Table 2). The S4 PGS method showed the greatest advancement in prediction accuracy and association for prostate cancer (AUC=0.710, OR=0.816) and schizophrenia (AUC=0.723, OR=0.844) than PRS-CS (prostate cancer AUC=0.684, OR=0.704; schizophrenia AUC=0.703, OR=0.778). Unlike S4 PGS which used fewer SNPs while varying across phenotypes, PRS-CS models steadily used 1.1 million SNPs per model. In brief, our results confirmed that S4 PGS methods outperformed LDPred2 and PRS-CS in predicting polygenic risk scores on multiple phenotypes.

### S4 PGS performance on various SNP selection thresholds

S4 PGS is a parsimonious model which uses the most significant SNPs and results in the selection of fewer SNPs. We investigated the influence of SNP selection thresholds on S4 PGS predictions. The threshold was determined by the measure of SNP p-value divided by squared correlation of linkage disequilibrium. We examined thresholds of 0.02, 0.15, and 0.6, which on average used 54,000, 253,000, 731,000 SNPs. For body mass index, rather than the 0.6 threshold, the S4 PGS model was tested with all SNPs, as the number of SNPs was still sufficiently small for the model to run. In general, the inclusion of more SNPs led to better prediction accuracy and association (Table 3). However, this comes at the penalty of larger SNP datasets and the need for increasing computational time. For example, the AUC values for body mass index are 0.27, 0.3, 0.31 for the three thresholds respectively, and the normalized ORs are 1.29, 1.42, and 1.47. The only exception is type 1 diabetes, which showed no improvement when increasing the threshold from 0.02 to 0.15. There was little difference in prediction performances when the threshold was increased from 0.15 to 0.6, while the computation time largely increased and models for breast cancer, endometrial cancer, prostate cancer, and type 1 diabetes became computationally infeasible. Even though the various number of SNPs selected also depends on the density of summary statistics coverage and shrinkage parameters of effect estimates, we confirmed through these threshold analyses that the use of threshold 0.15 in this study is ideal in balancing between model performance and computational time.

### External validations of S4 PGS in Finnish and Japanese populations

To assess the transferability of S4 PGS method to other populations we leveraged the existing GWAS summary statistics from FinnGen (Kurki *et al*., 2022) and BioBank Japan (Nagai *et al*., 2017) to examine the S4 PGS model performance. Some variants did not have summary statistics in the GWAS summary statistics, so the PGS was assessed using only the variants for which summary statistics were available, and this may have impacted the performance of the model in these cohorts. In particular, several variants were missing from the summary statistics that would have improved the type 1 diabetes result. The best fit model on the whole UK Biobank data for each phenotype was tested on the FinnGen and Biobank Japan populations. In most of the phenotypes the PGS associations from Finnish and Japanese ancestries are both comparable to the PGS associations obtained from the European cohort (Table 4). Larger ORs were reported in the FinnGen cohort than UK Biobank for breast cancer (0.611 vs 0.577), major depressive disorder (0.229 vs 0.224), and prostate cancer (0.858 vs 0.816). There was no phenotype in which a larger OR was calculated in BioBank Japan than UK Biobank (0.93. We speculate that the less transferability from European ancestry to Japanese ancestry might be attributed to their lower correlated genetic architectures.

We further compared the transferability of LDPred2 using the same evaluation metrics. The results were similar to those generated using the S4 model, with S4 performing better in phenotypes with stronger main effects (such as breast cancer and prostate cancer) and LDPred2 performing better in phenotypes where a very large number of variants contribute (such as MDD). When accessing the cross-biobank performance of both methods, the polygenic predictions of type 1 diabetes in the Japanese population are lower than expected for both S4 PGS (OR=0.068) and LDPred2 (OR=0.039). When examining the summary statistics, we discovered that top SNPs for type 1 diabetes in the European population were not significant in the Japanese population (Supplementary Table 3), explaining the discordance. In addition, height and body mass index reported much lower beta coefficients for the Japanese population (0.213 for height, and 0.186 for body mass index) than the European populations (1.468 for height, and 3.494 for body mass index), even though the standard errors of the estimates are much less when comparing to the European samples. This observation might be a result of variance in the traits in different populations.

Genotype level data were not available for FinnGen and Biobank Japan, so we evaluated the S4 PGS and LDPred2 predictive performance by estimating PGS effects (joint effects) from GWAS summary statistics (marginal effects) and variants correlation matrix (See Methods). To validate the rationality of this estimation approach, we compared S4 PGS and LDPred2 model results evaluated by directly calculating PGS with results assessed by estimating through summary statistics (Supplementary Table 4). The results from both approaches were similar, reinforcing the validity of our reported results in cross-biobank analyses. The chi-squared statistics, ORs, and confidence intervals show little discrepancy between the two approaches in all phenotypes for both S4 PGS and LDPred2. In particular, when S4 PGS overestimated the effect, LDPred2 tended to overestimate the effect and vice-versa. The only unfavorable agreement was observed in type 1 diabetes, where the OR is 1.073 when directly calculating PGS and 1.270 when estimating from summary statistics. Considering the PGS effect was dominated by the most significant SNPs, this might explain the observed difference. Taken together, we demonstrated the utility of S4 PGS in multi-phenotype cross-population risk predictions.

## Discussion

Genetic risk profiling with PGS can be used to stratify individuals according to their disease risks and could be used to improve screening and prevention strategies and reduce disease mortality (Khera *et al*., 2018; Mavaddat *et al*., 2019). Previously, we had demonstrated the improvement of the S4 PGS method over existing methods in predicting epithelial ovarian cancer risk. Here, we extended the S4 PGS method to 12 phenotypes in UK Biobank, and performed a systematic comparison with LDPred2 and PRS-CS. The S4 PGS method demonstrated improved AUCs and outperformed the other two models across multiple phenotypes. We assessed the effect of the number of SNPs included in the model on S4 PGS predictive performance by changing the SNP selection threshold. We identified a computationally efficient while accurate threshold, which could be used to guide parameter settings. Furthermore, we explored the transferability of S4 PGS in modeling joint SNP effects for risk prediction in Finnish and Japanese populations and compared them with common approaches. Our results provided stronger associations with risks of each phenotype.

The UK Biobank and other population-scale biobanks represent a useful resource for testing PGS models. As we have done here, comparing a particular PGS formation method to others across a variety of phenotypes and a variety of ancestries serves as a powerful benchmark of PGS model performance. Recently, a UK Biobank Polygenic Risk Score (PRS) method has been released as a resource of polygenic scores across many diseases and traits, with benchmarking of multiple PGS algorithms or published PGSs (Thompson *et al*., 2022) against this new method. Notably ovarian cancer was the only phenotype where the UKB generated PRS did not improve on the previously reported PRS, which we previously generated using the S4 method (Dareng et al. 2022). As population-scale biobanks continue to become available and grow, this benchmarking and comparison of different methods is helpful for developing and improving PGSs.

The S4 PGS model is complementary to LDPred2, which has been used widely to predict risks of polygenic traits. The two methods mainly differ in three aspects, which we address in detail below: The type of prior on SNP effect sizes, correlation matrix computation, and SNP selection. The S4 PGS method places a continuous shrinkage prior on SNP effect sizes, and LDPred2 uses the common default spike-and-slab prior. The continuous shrinkage prior can model distributions with heavy tails better, whereas it can be more vulnerable if there are inaccuracies in the reference correlation matrix. To reduce the time of computing the variant correlation matrix, S4 PGS partitions SNPs into blocks that are roughly independent of each other, and performs SNP selection for each block. LDPred2 assumes a sparse matrix where for a given SNP, the correlations with other SNPs are set to zero if the genetic distance is greater than 3 centimorgan. The S4 approach reduces computational burden and still maintains accuracy when SNPs are reasonably independent of each other or have only minor effects. Lastly, the S4 method considers all SNPs and selects them based on ranked position by P value, (i.e. the most significant first) that are not excessively correlated with already selected SNPs. This ensures a parsimonious model that requires fewer SNPs. On the other hand, LDPred2 focuses only on the Hapmap3 SNPs, which are better imputed and can be applied in all PGS models.

Further optimization of the S4 PGS models could be achieved by examining the model parameters in greater detail. There are a number of parameters used to generate the models, as well as the core continuous shrinkage prior parameters. In this study, we primarily assessed the effect of SNPs selection threshold. Parameters such as correlation threshold for adding SNPs into the model, maximum individual correlation, and quality control criteria for summary statistics were set based on our prior experience (Dareng *et al*., 2022). A systematic tuning of these thresholds by phenotype may increase the robustness of S4 PGS models.

In conclusion, our results indicate that S4 PGS provides improvements in risk prediction for multiple phenotypes over more common approaches. Our approach overcomes the computational limitations without loss of accuracy. Besides, S4 PGS provides sufficient evidence of transferability to populations of other ancestries. Future works can be focused on the incorporation of epidemiological risk factors or SNPs functional annotations, to further improve the predictive power.

## Data Availability

All data produced in the present work are contained in the manuscript.

https://github.com/jpt34/S4_programs

## Data Availability Statement

The S4 program is freely available at https://github.com/jpt34/S4_programs.

