## Supplemental Tables for "Improving on polygenic scores across complex traits using select and shrink with summary statistics"

**Supplementary Table 1. Summary statistics from UK Biobank, FinnGen and Biobank Japan**

| **Phenotype** | **GWAS citation** | **Sample size (case/control)** | **Number of variants** |
| --- | --- | --- | --- |
| **UK Biobank** |  |  |  |
| Asthma | [PMID:[29273806](https://www.ncbi.nlm.nih.gov/pubmed/29273806)] | 19954 / 107715 | 2001280 |
| Body mass index | [PMID:[25673413](https://www.ncbi.nlm.nih.gov/pubmed/25673413)] | 322000 * | 2529672 |
| Breast cancer | [PMID:[29059683](https://www.ncbi.nlm.nih.gov/pubmed/29059683)] | 137045 / 119078 | 11792542 |
| Coronary artery disease | [PMID:[26343387](https://www.ncbi.nlm.nih.gov/pubmed/26343387)] | 60801 / 123504 | 9455778 |
| Endometrial cancer | [PMID:[27008869](https://www.ncbi.nlm.nih.gov/pubmed/27008869)] | 12906 / 108979 | 9529047 |
| Height | [PMID:[25282103](https://www.ncbi.nlm.nih.gov/pubmed/25282103)] | 253000 * | 2550839 |
| Inflammatory bowel disease | [PMID:[28067908](https://www.ncbi.nlm.nih.gov/pubmed/28067908)] | 25042 / 34915 | 9584779 |
| Major depressive disorder | [PMID:[29700475](https://www.ncbi.nlm.nih.gov/pubmed/29700475)] | 59851 / 113154 ** | 13554550 |
| Prostate cancer | [PMID:[29892016](https://www.ncbi.nlm.nih.gov/pubmed/29892016)] | 79148 / 61106 | 20370946 |
| Schizophrenia | [PMID:[21926974](https://www.ncbi.nlm.nih.gov/pubmed/21926974)] | 59851 / 113154 ** | 11308214 |
| Type 1 diabetes | [PMID:[28763444](https://www.ncbi.nlm.nih.gov/pubmed/28763444)] | 5913 / 8828 | 9038020 (4189165) *** |
| Type 2 diabetes | [PMID:[28566273](https://www.ncbi.nlm.nih.gov/pubmed/28566273)] | 26676 / 132532 | 12056346 (10607793) *** |
| **FinnGen** | **Phenotype Code** |  |  |
| Asthma | J10_ASTHMA | 25544 / 158452 | 16380175 |
| Breast cancer | C3_BREAST | 11573 / 135488 | 16379783 |
| Cardiovascular disease^a^ | I9_CORATHER | 28598 / 222551 | 16380465 |
| Endometrial cancer | C3_CORPUS_UTERI | 1430 / 145631 | 16379783 |
| Inflammatory bowel disease | K11_IBD | 7206 / 253199 | 16380465 |
| Major depressive disorder | F5_DEPRESSIO | 28098 / 228817 | 16380456 |
| Prostate cancer | C3_PROSTATE | 8709 / 104635 | 16378834 |
| Schizophrenia | F5_SCHZPHR | 5760 / 249610 | 16380455 |
| Type 1 diabetes | E4_DM1 | 7609 / 215160 | 16380007 |
| Type 2 diabetes | E4_DM2 | 41245 / 215160 | 16380439 |
| **BioBank Japan** |  |  |  |
| Asthma |  | 13015 / 162933 | 13435616 |
| Body mass index |  | 163835 * | 13236464 |
| Breast cancer |  | 6325 / 73225 | 13407080 |
| Endometrial cancer |  | 1200 / 60614 | 13387811 |
| Height |  | 165056 * | 13236464 |
| Major depressive disorder |  | 836 / 177794 | 13436071 |
| Prostate cancer |  | 5672 / 84660 | 13412985 |
| Schizophrenia |  | 99 / 177794 | 13435901 |
| Type 1 diabetes |  | 1219 / 132032 | 13427790 |
| Type 2 diabetes |  | 45383 / 132032 | 13435872 |

**Supplementary Table 2. Comparison between S4 PGS and PRS-CS with 10-fold cross-validation on multiple phenotypes.** AUC values with better performance are highlighted in bold.

| **Phenotype** | **S4** | | | **PRS-CS** | | |
| --- | --- | --- | --- | --- | --- | --- |
|  | **Number of SNPs** | **AUC** | **OR** | **Number of SNPs** | **AUC** | **OR** |
| Asthma | 425752 | 0.600 | 0.362 | 986459 | 0.587 | 0.305 |
| Body mass index | 585796 | **0.311** | 1.468 | 1023391 | 0.302 | 1.426 |
| Breast cancer | 357287 | **0.658** | 0.579 | 1119952 | 0.646 | 0.536 |
| Coronary artery disease | 689356 | 0.648 | 0.585 | 1116009 | 0.641 | 0.557 |
| Endometrial cancer | 271090 | **0.607** | 0.396 | 1112889 | 0.594 | 0.344 |
| Height | 461803 | **0.368** | 3.494 | 1021155 | 0.356 | 3.361 |
| Inflammatory bowel disease | 358464 | **0.725** | 0.857 | 1085126 | 0.707 | 0.778 |
| Major depressive disorder | 843583 | 0.563 | 0.224 | 1119734 | 0.560 | 0.215 |
| Prostate cancer | 409227 | **0.710** | 0.816 | 1120676 | 0.683 | 0.704 |
| Schizophrenia | 1239271 | **0.723** | 0.844 | 1119339 | 0.700 | 0.767 |
| Type 1 diabetes | 19584 | 0.793 | 1.149 | 516188 | 0.752 | 0.890 |
| Type 2 diabetes | 874431 | **0.662** | 0.608 | 1119580 | 0.654 | 0.570 |

OR: log odds ratio per 1 standard deviation of PGS

**Supplementary Table 3. Top hits for type 1 diabetes compared between UK Biobank and BioBank Japan.**

| **SNP** | **Chr** | **Position** | **Effect** | **Baseline** | **UK Biobank** | | | | **Biobank Japan** | | | |
| --- | --- | --- | --- | --- | --- | --- | --- | --- | --- | --- | --- | --- |
|  |  |  |  |  | **EAF** | **OR** | **SE** | **P-value** | **EAF** | **OR** | **SE** | **P-value** |
| rs10496344 | 2 | 100764125 | C | T | 0.38 | -0.145 | 0.026 | 2.45E-08 | 0.470 | 0.049 | 0.019 | 0.011 |
| rs33998987 | 2 | 163284067 | C | T | 0.336 | -0.198 | 0.028 | 1.53E-12 | 0.683 | -0.025 | 0.021 | 0.230 |
| rs3087243 | 2 | 204738919 | A | G | 0.47 | -0.178 | 0.025 | 1.08E-12 | 0.273 | 0.006 | 0.021 | 0.781 |
| rs11727369 | 4 | 123286227 | A | G | 0.377 | 0.161 | 0.025 | 1.19E-10 | 0.627 | -0.035 | 0.020 | 0.079 |
| rs9366622 | 6 | 25414537 | C | T | 0.862 | -0.327 | 0.034 | 6.74E-22 | 0.984 | 0.054 | 0.077 | 0.482 |
| rs3129783 | 6 | 32655730 | G | A | 0.561 | 1.923 | 0.041 | 0 | 0.574 | 0.003 | 0.020 | 0.862 |
| rs4142967 | 6 | 90996349 | T | C | 0.459 | 0.155 | 0.025 | 5.65E-10 | 0.030 | -0.100 | 0.057 | 0.078 |
| rs7034200 | 9 | 4289050 | A | C | 0.479 | 0.145 | 0.025 | 6.63E-09 | 0.413 | 0.015 | 0.019 | 0.442 |
| rs7931848 | 11 | 2270029 | A | C | 0.113 | -0.253 | 0.042 | 1.70E-09 | 0.044 | -0.037 | 0.047 | 0.428 |
| rs7297175 | 12 | 56473808 | C | T | 0.588 | -0.246 | 0.025 | 7.57E-23 | 0.802 | 0.003 | 0.024 | 0.913 |
| rs55993634 | 16 | 75236763 | G | C | 0.087 | 0.294 | 0.045 | 6.43E-11 | 0.243 | 0.020 | 0.023 | 0.393 |
| rs2186941 | 18 | 12772622 | T | C | 0.218 | 0.186 | 0.03 | 5.65E-10 | 0.211 | -0.008 | 0.025 | 0.743 |
| rs6043405 | 20 | 1615544 | C | T | 0.662 | 0.144 | 0.026 | 3.05E-08 | 0.841 | 0.005 | 0.026 | 0.858 |
| rs80054410 | 21 | 43836010 | C | T | 0.357 | 0.175 | 0.025 | 2.56E-12 | 0.040 | -0.085 | 0.049 | 0.087 |
| rs5763790 | 22 | 30522413 | C | G | 0.616 | -0.166 | 0.025 | 3.14E-11 | 0.755 | -0.013 | 0.023 | 0.576 |

EAF: effect allele frequency; OR: odds ratio; SE: standard deviation

**Supplementary Table 4. Comparisons of S4 and LDPred models performance evaluated by directly calculating PGS and by estimating through summary statistics.** The multiple phenotypes were collected from UK BioBank.

| **Phenotype** | **Directly calculating PGS** | | | **Comparing summary statistics** | | |
| --- | --- | --- | --- | --- | --- | --- |
|  | **χ^2^** | **OR** | **95% CI** | **χ^2^** | **OR** | **95% CI** |
| **S4** |  |  |  |  |  |  |
| Asthma | 5388 | 0.362 | 0.353-0.372 | 5448 | 0.358 | 0.348-0.367 |
| Body mass index | 43473 | 1.468 | 1.455-1.482 | 39809 | 1.476 | 1.461-1.490 |
| Breast cancer | 1467 | 0.577 | 0.547-0.607 | 1448 | 0.560 | 0.531-0.589 |
| Coronary artery disease | 3225 | 0.585 | 0.564-0.605 | 3228 | 0.571 | 0.551-0.591 |
| Endometrial cancer | 171 | 0.406 | 0.345-0.467 | 190 | 0.404 | 0.345-0.462 |
| Height | 169086 | 3.494 | 3.478-3.511 | 125121 | 3.497 | 3.478-3.516 |
| Inflammatory bowel disease | 2493 | 0.856 | 0.822-0.89 | 2588 | 0.855 | 0.822-0.888 |
| Major depressive disorder | 778 | 0.224 | 0.208-0.24 | 823 | 0.223 | 0.208-0.239 |
| Prostate cancer | 3030 | 0.816 | 0.786-0.846 | 2824 | 0.766 | 0.738-0.794 |
| Schizophrenia | 305 | 0.858 | 0.762-0.955 | 339 | 0.840 | 0.749-0.930 |
| Type 1 diabetes | 603 | 1.140 | 1.049-1.231 | 723 | 1.270 | 1.178-1.363 |
| Type 2 diabetes | 1167 | 0.609 | 0.574-0.645 | 1174 | 0.592 | 0.558-0.626 |
| **LDPred2** |  |  |  |  |  |  |
| Asthma | 5498 | 0.366 | 0.356-0.376 | 5598 | 0.361 | 0.352-0.371 |
| Body mass index | 43338 | 1.466 | 1.452-1.480 | 39421 | 1.479 | 1.465-1.494 |
| Breast cancer | 1388 | 0.562 | 0.532-0.592 | 1383 | 0.546 | 0.517-0.575 |
| Coronary artery disease | 3101 | 0.579 | 0.558-0.600 | 3049 | 0.558 | 0.539-0.578 |
| Endometrial cancer | 128 | 0.352 | 0.291-0.413 | 141 | 0.351 | 0.292-0.409 |
| Height | 172261 | 3.502 | 3.486-3.519 | 126737 | 3.532 | 3.513-3.551 |
| Inflammatory bowel disease | 2310 | 0.830 | 0.796-0.864 | 2389 | 0.820 | 0.787-0.852 |
| Major depressive disorder | 800 | 0.227 | 0.212-0.243 | 828 | 0.228 | 0.212-0.243 |
| Prostate cancer | 2596 | 0.755 | 0.725-0.784 | 2476 | 0.716 | 0.688-0.744 |
| Schizophrenia | 288 | 0.831 | 0.735-0.928 | 311 | 0.818 | 0.728-0.909 |
| Type 1 diabetes | 570 | 1.073 | 0.986-1.160 | 692 | 1.249 | 1.157-1.341 |
| Type 2 diabetes | 1123 | 0.596 | 0.561-0.631 | 1130 | 0.583 | 0.549-0.618 |
